# Addressing inter-individual variability in CSF levels of brain-derived proteins across neurodegenerative diseases

**DOI:** 10.1101/2024.05.27.24307739

**Authors:** Sára Mravinacová, Sofia Bergström, Jennie Olofsson, Nerea Gómez de San José, Sarah Anderl-Straub, Janine Diehl-Schmid, Klaus Fassbender, Klaus Fliessbach, Holger Jahn, Johannes Kornhuber, G. Bernhard Landwehrmeyer, Martin Lauer, Johannes Levin, Albert C. Ludolph, Johannes Prudlo, Anja Schneider, Matthias L. Schroeter, Jens Wiltfang, Petra Steinacker, Markus Otto, Peter Nilsson, Anna Månberg, FTLD consortium

**Affiliations:** Department of Protein Science, KTH Royal Institute of Technology, SciLifeLab, Stockholm, Sweden; Department of Neurology, University Hospital Ulm (UKU), Ulm, Germany; Department of Psychiatry, Technical University of Munich, Munich, Germany; kbo-Inn-Salzach-Klinikum gemeinnützige GmbH, Wasserburg am Inn, Germany; Department of Neurology, Saarland University, Homburg, Germany; Department of Neurodegenerative Diseases and Geriatric Psychiatry, University of Bonn and DZNE Bonn, Bonn, Germany; Department of Psychiatry, University Hospital Hamburg, Germany; Department of Psychiatry, Friedrich-Alexander University Erlangen-Nuremberg, Erlangen, Germany; Center for Mental Health, Department of Psychiatry, Psychosomatics and Psychotherapy, University Hospital Würzburg, Würzburg, Germany; Department of Neurology, LMU University Hospital, LMU Munich, Munich, Germany; German Center for Neurodegenerative Diseases, site Munich, Munich, Germany; Munich Cluster for Systems Neurology (SyNergy), Munich, Germany; German Center for Neurodegenerative Diseases (DZNE e.V.), Ulm, Germany; Rostock University Medical Center and German Center for Neurodegenerative Diseases (DZNE), Rostock, Germany; Clinic for Cognitive Neurology, University Clinic Leipzig, and Max Planck Institute for Human Cognitive and Brain Sciences, Leipzig, Germany; Department of Psychiatry and Psychotherapy, University Medical Center Goettingen, and DZNE, Goettingen, Germany; Neurosciences and Signaling Group, Institute of Biomedicine (iBiMED), Department of Medical Sciences, University of Aveiro, Aveiro, Portugal; Department of Neurology, Martin-Luther-University Halle-Wittenberg, Halle (Saale), Germany

## Abstract

**Background:** Accurate diagnosis and monitoring of neurodegenerative diseases requires reliable biomarkers. Cerebrospinal fluid (CSF) proteins hold promise for reflecting brain pathology, yet their utility may be compromised by natural variability between individuals, weakening their association with disease and impacting their diagnostic accuracy.

**Method:** We measured the levels of 69 proteins in CSF from 499 individuals using an antibody-based suspension bead array technology. In this multi-disease cohort including participants with Alzheimer’s disease, behavioural variant frontotemporal dementia, primary progressive aphasias, amyotrophic lateral sclerosis, corticobasal syndrome, primary supranuclear palsy and healthy controls, we investigated inter-individual variability in CSF protein levels. Further, we explored protein profiles across the diseases, after adjusting for this variability using linear modelling.

**Results:** Correlation and PCA analysis on the measured CSF proteins revealed the presence of inter-individual variability in overall CSF levels of brain-derived proteins, which could not be explained by disease associations. Using linear modelling, we show that adjusting for median CSF levels of brain-derived proteins increases the diagnostic accuracy of many proteins, including those previously identified as altered in CSF in the context of neurodegenerative disorders. We further demonstrate a simplified approach for the adjustment using pairs of correlated proteins with opposite alteration in the diseases. With this approach, the proteins adjust for each other and further increase the biomarker performance through additive effect. Moreover, we find that many CSF proteins show similar alteration across the studied diseases, indicating that these proteins likely reflect shared processes.

**Conclusion:** CSF protein profiles are affected by inter-individual variability which partially hinders the diagnostic potential of CSF proteins. Thus, we propose that this variability should be accounted for in future studies. Further, we demonstrate high overlap in CSF protein patterns across neurodegenerative diseases, highlighting the need for multi-disease studies in order to identify biomarkers that are specific to or common between different diseases.

## Introduction

Neurodegenerative disorders encompass a variety of diseases characterised by a gradual loss of neurons. The continuously increasing prevalence, associated burden of deaths, and rising costs underscore the recognition of neurodegenerative disorders as a major public health challenge [1]. Dementias represent a large proportion of these diseases, with Alzheimer’s disease (AD) being the most common form. The diagnosis of neurodegenerative disorders, and especially the different dementia forms, is cumbersome due to the large overlap of clinical symptoms [2, 3]. Consequently, the need for specific and reliable biomarkers is growing, not only to identify the disease, but also for monitoring the disease progress and to evaluate the efficiency of new potential treatments. Further, biomarker research can provide better understanding of the pathological processes underlying the disease mechanisms on a molecular level.

A cost effective and relatively non-invasive approach to monitor pathological processes involves measuring protein levels in biofluid samples. Although blood-derived samples are less invasive, cerebrospinal fluid is considered a more informative sample type due to its close proximity to the brain tissues [4]. Amyloid beta (Aβ) 1-42 peptide and phosphorylated tau (p-tau) are cerebrospinal fluid (CSF) biomarker indicative of the presence of Aβ plaques and tau-neurofibrillary tangles in the brain tissue, the hallmarks of AD. These biomarkers are now broadly used to aid in AD diagnosis as well as to select participants for clinical studies [5].

Although the number of protein biomarker studies has been steeply increasing in recent years, only a handful of biomarkers have been implemented in clinical practise, partially due to the challenges associated with biomarker research and validation [6]. One of these challenges is the presence of inter-individual variability in CSF protein levels unrelated to disease, which can lead to low biomarker performance [7, 8]. Although the sources of such variability are complex, indirect adjustments can be made to reduce its impact. A good example used in practice is the CSF Aβ42 in ratio with Aβ40, where Aβ40 works as an adjustment factor, increasing the accuracy of Aβ42 as a predictor for the presence of amyloid plaques in the brain [9, 10]. Interestingly, several studies, including ours, revealed relatively strong correlations between Aβ40 and p-tau [11, 12] as well as between other CSF brain-derived proteins across various neurodegenerative diseases and in healthy individuals [11, 13-19]. This broad co-variance pattern among CSF proteins may indicate possible differences in overall CSF protein levels between individuals, which might be driving the primary variance in CSF protein levels independently of pathology.

We have previously demonstrated that adjusting for overall CSF protein levels by using CSF proteins in ratios enhances the performance of biomarkers within AD [19]. Building upon this work, we expand our research to a spectrum of neurodegenerative diseases including AD, amyotrophic lateral sclerosis (ALS), behavioural variant frontotemporal dementia (bvFTD), corticobasal syndrome (CBS), primary progressive aphasia (PPA), and progressive supranuclear palsy (PSP). Analysing a panel of 69 proteins in CSF, pre-selected based on previous in-house studies and literature, we initially explore the overall variability in the dataset and confirm the presence of inter-individual variability in general levels of brain-derived CSF proteins. Subsequently, we evaluate different approaches to adjust for this variability in order to improve biomarker performance. Importantly, we compare the observed CSF profiles across the included diseases to discern differences and similarities, providing valuable insights into disease-specific and unspecific protein signatures.

## Materials and methods

### Participants

The 499 participants included patients with Alzheimer’
ss disease (AD) (n=69), primary progessive aphasias (PPA) (n=166), behavioural frontotemporal demenia (bvFTD) (n=129), corticobasal syndrome (CBS) (n=26), progressive supranuclear palsy (PSP) (n=39), amyotrophic lateral sclerosis (ALS) (n=35), and healthy controls (n=35) (Table 1). Diagnosis was based on internationally established criteria. Participants were examined between 05/2011 and 07/2020 within the FTLD Consortium study [20]. Each participant passed through comprehensive clinical and neuropsychological tests, as well as CSF and blood sampling and MRI. CSF was obtained by lumbar puncture. The FTLD Consortium study applies strict standard operating procedures (SOP) to guarantee data reliability. The study was conducted following the Declaration of Helsinki [21] and approved by the local ethics committees of all centers involved (University of Ulm #39/11). Participants gave written informed consent.

**Table 1:** Cohort description.

|  | AD | ALS | bvFTD | CBS | PPA | PSP | Healthy control |
| --- | --- | --- | --- | --- | --- | --- | --- |
| **N** | 69 | 35 | 129 | 26 | 166 | 39 | 35 |
| **Female [N](%)** | 40 (58%) | 15 (43%) | 48 (37%) | 14 (54%) | 77 (46%) | 20 (51%) | 19 (54%) |
| **Age<sup>a</sup>** | 70 (47-83) | 66 (42-85) | 62 (41-82) | 69 (54-82) | 68 (44-86) | 68 (50-85) | 73 (50-89) |
| **Aβ42<sup>a</sup>** | 498 (202-1415) | 897 (393-1828) | 784 (251-1950) | 736 (252-1461) | 776 (210-2087) | 661 (318-1445) | - |
| **Aβ40<sup>a</sup>** | 9789 (783-20 010) | 10 516 (4627-20 301) | 9112 (2756-20 158) | 9710 (3000-13 221) | 10 780 (2948-21 447) | 8719 (3787-17 438) | - |
| **Aβ42/40<sup>a</sup>** | 0.046 (0.029-0.185) | 0.093 (0.029-0.118) | 0.095 (0.036-0.122) | 0.086 (0.045-0.116) | 0.091 (0.024-0.123) | 0.093 (0.030-0.116) | - |
| **t-tau<sup>a</sup>** | 638 (170-2000) | 376 (145-953) | 334 (88-2,000) | 354 (98-1109) | 402 (82-2,000) | 284 (110-1,672) | - |
| **p-tau<sup>a</sup>** | 106 (12-352) | 36 (15-134) | 40 (10-378) | 46 (14-164) | 48 (10-381) | 40 (16-253) | - |
| **Q-alb<sup>b</sup>** | 6.09 (2.80-14.60) | 6.40 (2.80-18.20) | 6.10 (2.60-23.50) | 5.37 (2.70-17.00) | 6.20 (2.70-14.20) | 5.80 (2.90-24.60) | - |
| **CDR score<sup>a,b,c</sup>** | 5.2 (0.5-14.0) | 2.8 (0.0-17.0) | 5.0 (0.0-17.0) | 4.0 (0.0-11.0) | 2.5 (0.0-16.0) | 3.8 (0.0-18.0) | - |

<sup>a</sup> Data is presented in the format: median (range).
<sup>b</sup> Q-Alb – missing data [N]: AD: 19 (28%), ALS: 4 (11%), bvFTD: 29 (22%), CBS: 3 (12 %), PPA: 53 (32%), PSP: 8 (21%).
<sup>c</sup> CDR score – missing data [N]: AD: 11 (16%), ALS: 7 (20%), bvFTD: 17 (13%), CBS: 6 (23 %), PPA: 27 (16%), PSP: 9 (23 %).

### CSF amyloid and tau measurements

Quantification of CSF Aβ40, Aβ42 and p-tau181 was conducted using the chemiluminescent enzyme immunoassay on the on the automated Lumipulse G600II analyzer according to the instructions (Fujirebio). Albumin in CSF and serum was measured by standard immunochemical nephelometry [22, 23]. To analyze the association between CSF AD markers and median CSF protein levels, individuals were classified based on their CSF amyloid and tau status, independent of diagnosis. Individuals with Aβ42/40 ≤ 0.07 were classified as amyloid positive (A+) and with p-tau ≥ 60 as tau positive (T+), resulting into four groups, A-T- (n = 258), A+T- (n = 30), A+T+ (n = 142) and A-T+ (n = 34).

### CSF protein analysis using suspension bead array

The selection of proteins to include in this study was mainly based on prior in-house studies [11, 13, 24-28]. The pre-selected proteins were analysed using a multiplex antibody-based suspension bead array employing polyclonal antibodies produced in rabbits within the Human Protein Atlas project (www.proteinatlas.org). Each antibody targeting a selected protein was attached to the surface of color-coded magnetic beads (MagPlex, Luminex corp.) through NHS-EDC chemistry as previously described by Pin *et al*. [29]. The antibody-coupled beads were pooled into two separate bead arrays, each containing bead-IDs corresponding to the intended protein target analysis at different sample dilutions (either 1/25 or 1/200) (**Supp. Table 1**).

The CSF samples were distributed into 96-well PCR plates in a stratified randomisation manner based on diagnosis, age and sex of the individuals as well as the sample collection site. The crude samples were then directly labelled with biotin (NHS-PEG4-biotin, A39259, ThermoFisher Scientific) at an approximate tenfold molar excess, following established procedures [29]. The biotin-labelled samples were diluted with a buffer containing bovine serum albumin (BSA) and rabbit IgG to a final dilution of 1/25 and 1/200. The diluted samples were further heat-treated for 30 min at 56 °C before incubation with the prepared bead arrays at room temperature overnight. Following the washing of unbound proteins, the antibody-bound protein targets were incubated with a streptavidin-coupled fluorophore. The signal read-out was facilitated using a Flexmap 3D instrument (Luminex corp.). The data per bead ID (protein target) and sample was acquired as a median fluorescent intensity, yielding relative protein quantities. The obtained data were further adjusted for delayed instrument readout using robust linear model as described previously [19], and for sample plate batch-effect using removebatchEffect (limma). To evaluate the intra-assay variability of each protein assay, three sample pool replicates were included in each 96-well sample plate. Further, one 96-well sample plate was re-analyzed to evaluate inter-assay correlation. Data analysis was conducted on 69 proteins which passed the quality control. (**Supp. Table 1**).

### Data analysis and visualizations

The open-source R statistical software (version 4.2.2) was used for all data processing, analysis, and visualization, incorporating supplementary packages tidyverse, tidymodels, ggpubr, ggbeeswarm, ggrepel, pheatmap, stats, scales, and patchwork. Details of additional packages and functions employed in specific data analysis sections are provided therein. To enhance the figure clarity and readability (e.g., to adjust figure legends) we performed additional refinements using the vector graphic editor Affinity Designer (version 1.8.6) by Serif, West Bridgford, UK.

For all analysis, relative protein levels measured using the suspension bead array were log2 transformed and centred to median (per protein). Correlation between the measured proteins was calculated on the full sample cohort using Spearman’s correlation (cor, stats). Hierarchical clustering in heatmaps was performed using Ward’s method (ward.D2) with Euclidean distance as the similarity measure. Whether proteins were classified as brain-elevated was determined based on the tissue transcriptomic data classification of genes from the Human Protein Atlas [30]. Principal component analysis was performed on log2 transformed data centred to mean and scaled to standard deviation. The median CSF protein levels were determined by calculating the median level of cluster 1 proteins per sample. The differences in median CSF protein levels between males and females were evaluated using the Wilcoxon’s two-sided test, and between the diagnostic groups using the Kruskal-Wallis test followed by Dunn’s test for pairwise comparisons, with Benjamini-Hochberg adjustment for multiple testing [31]. Adjusted p-values <0.05 were considered significant.

#### Linear regression models

Generalized linear models (glm, stats) were used to evaluate whether adjustment for general protein levels estimated by the sample median of cluster 1 proteins increased association of the analysed CSF proteins to individual diseases/healthy state. For this, two types of models were constructed for each measured protein and each comparison of two different diagnosis. In each model, the measured CSF protein levels were used as an outcome variable. In the first model, diagnosis was used as the main predictor and in the second model both diagnosis and cluster 1 protein sample median were used as predictors. Both models were further adjusted for sex and age. To compare the association of CSF protein levels of the analysed protein to the tested diagnosis with and without adjustment to general protein levels, the beta coefficient for diagnosis along with the p-value were extracted from the model. The extracted p-value was further adjusted for multiple comparisons using the Benjamini-Hochberg method. Adjusted p-values < 0.05 were considered significant.

#### Logistic regression models

To predict diagnosis, logistic regression models (glm, stats) were constructed. Two types of diagnosis predictions were investigated in this study, distinguishing individual diseases from healthy controls and distinguishing individual diseases from all other diseases. For both cases, several different predictor combinations were tested: 1) only cluster 1 protein sample median, 2) only individual CSF proteins, 3) cluster 1 protein sample median + individual CSF proteins, 4) CSF protein pair combinations. Given the limited size of the sample groups, model performance was evaluated using cross-validation, with the area under the receiver operating characteristic (ROC) curve (AUC) as the primary metrics. Further, to address imbalances in sample group sizes, undersampling to the size of the smaller group was applied. Each model iteration involved 15 rounds of undersampling with 3 partitions and 5 repeats in cross-validation for each undersampling round. The resulting AUC represented the median AUC across the 15 undersampling rounds, reported with the corresponding interquartile range (IQR) in the example ROC-curve plots and tables. The ROC-curves for example models were constructed using average sample predictions from the undersampling round resulting in the median AUC.

## Results

### Median CSF levels of brain-derived proteins represent the main variability between individuals

Based on the protein-to-protein correlation within the full cohort of 499 individuals, the 69 measured CSF proteins clustered in three main groups (Fig. 1A). Cluster 1 predominantly comprised proteins of brain origin, with weak correlation with the albumin CSF/serum quotient. The second cluster (cluster 2) instead comprised proteins with strong correlation to albumin quotient, suggesting either their transfer to CSF from blood [32], or low detectability of these proteins in CSF. Both complement component 9 (C9) and inter-alpha-trypsin inhibitor heavy chain 1 (ITIH1), which were included as controls for blood-derived proteins, cluster to this group. Two proteins, neurofilament medium (NEFM) and Chitinase 1 (CHIT1) clustered separately (cluster 3) as they did not correlate well to proteins in the two other clusters. Principal component analysis (PCA) performed on highly-intercorrelated proteins included in the predominantly brain-derived cluster 1 further showed that over 70 % of the variability in the data could be explained by median protein levels calculated from the proteins included in the cluster per sample (Fig. 1B, C). That is, some individuals in the cohort had consistently low levels across these proteins while other individuals had consistently high levels.

**Figure 1:**
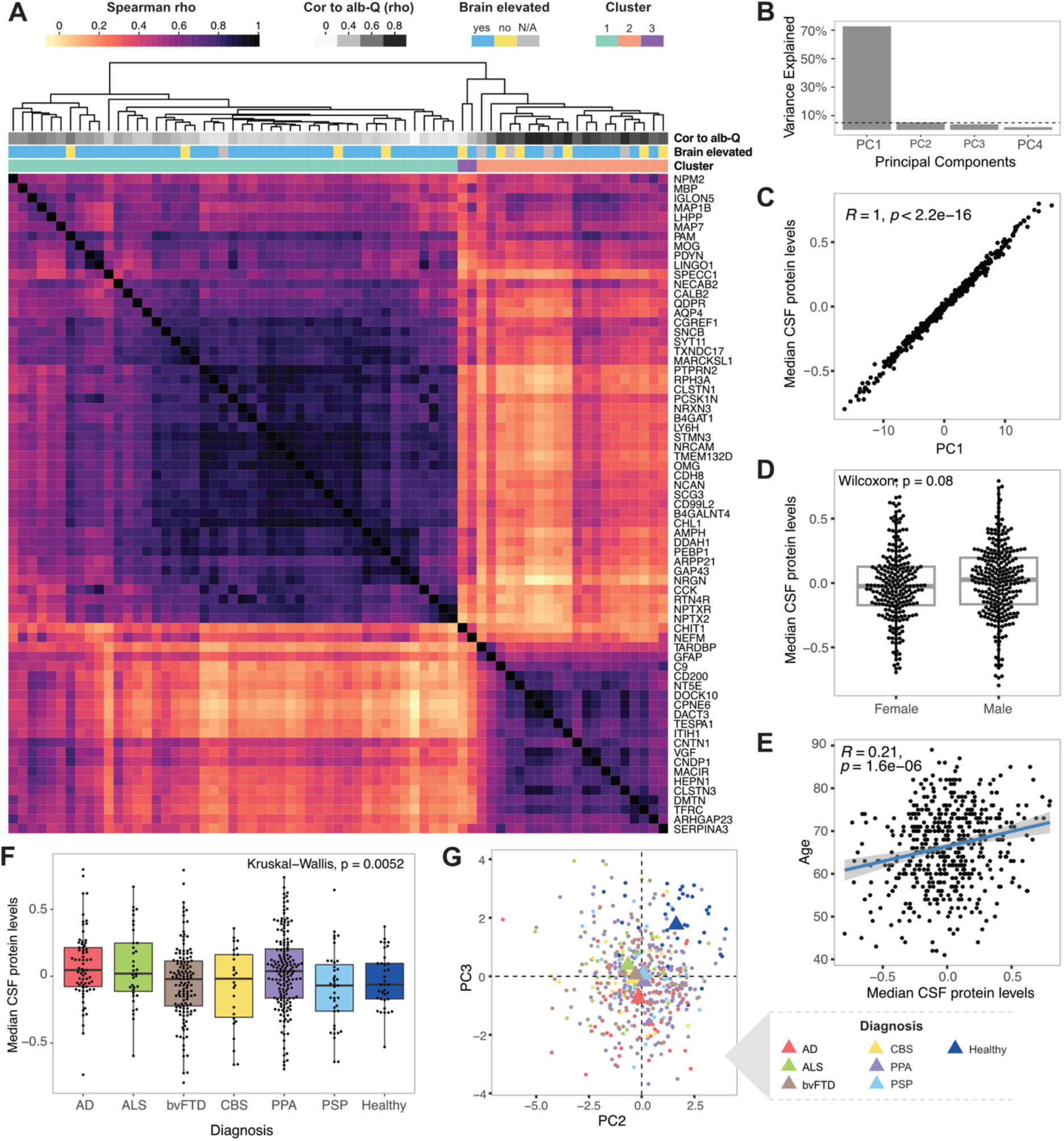
Median CSF protein levels account for the main variability in brain-derived proteins between samples. (**A**) A heatmap showing Spearman correlations between the measured proteins using the full sample cohort of 499 individuals. The heatmap is annotated with correlation of the individual proteins to albumin CSF/serum quotient, and classification of the proteins based on specificity to brain tissues according to the transcriptomic data provided in the Human Protein Atlas (www.proteinatlas.org). (**B**) Variance in the data explained by the first four principal components from principal component analysis (PCA) performed on all samples using cluster 1 proteins. (**C**) Pearson correlation between PC1 and median levels of cluster 1 proteins per individual. (**D**) Comparison of median CSF protein levels in males and females in the full cohort. (**E**) Pearson correlation between age and median CSF protein levels. (**F**) Comparison of median CSF protein levels between individuals stratified based on diagnosis. (**G**) Visualization of separation of healthy individuals from the rest of the cohort provided by PC2 and PC3.

Next, we investigated whether the differences in median levels of CSF brain-derived proteins (cluster 1) (further referred to as “median CSF protein levels”) could be associated with age, sex or diagnosis. While no significant differences were observed between the sexes (**Fig. 1D**), age demonstrated weak yet statistically significant positive correlation to median CSF protein levels (**Fig. 1E**). Subtle differences were also observed between the individual diagnostic groups (**Fig. 1F**). However, there was substantial overlap in the observed variance among the groups, and only bvFTD compared to AD demonstrated a statistically significant difference (**Supp. Table 2**). This suggests that factors other than sex, age or diagnosis primarily contributed to the variance in CSF median protein levels among individuals. Notably, while PC1 (∼ median CSF protein levels) did not show large variance between the studied diagnostic groups, PC2 and PC3 partially separated healthy individuals from those with neuro-degenerative diseases (**Fig. 1G**). However, PC2 and PC3 together only accounted for ∼ 10 % of the observed variance in the data (**Fig. 1B**). We hypothesise that adjusting the measured protein data for median CSF protein levels could decrease the influence of inter-individual variability in general protein levels, and potentially improve the diagnostic performance of the proteins.

**Table 2:** Protein specificity towards a single disease.

| Diagnosis | Protein | AUC (IQR) -<br>form other<br>diseases | AUC (IQR)<br>- from<br>healthy |
| --- | --- | --- | --- |
| AD | NRGN | 0.73<br>(0.72–0.74) | 0.82<br>(0.80–0.84) |
| ALS | NEFM | 0.77<br>(0.75–0.79) | 0.92<br>(0.91–0.92) |
| ALS | MBP | 0.72<br>(0.70–0.77) | 0.81<br>(0.80–0.81) |

### Adjusting for median CSF protein levels increases the performance of proteins in disease vs. healthy classification

To test whether adjustment for median CSF protein levels could enhance the association of measured proteins with disease, linear regression models were constructed for each protein and individual disease. We then compared p-values for diagnosis (in comparison to healthy controls) with and without adjustment for median CSF protein levels, while controlling for age and sex in all models. As presented in **Fig. 2A**, adjustment for the median CSF protein levels increased the association with individual diseases for the majority of the measured proteins. Particularly notable was the shift in statistical significance for proteins with decreased CSF protein levels in the individual diseases compared to healthy controls; a majority of these proteins only reached statistical significance after adjustment.

**Figure 2:**
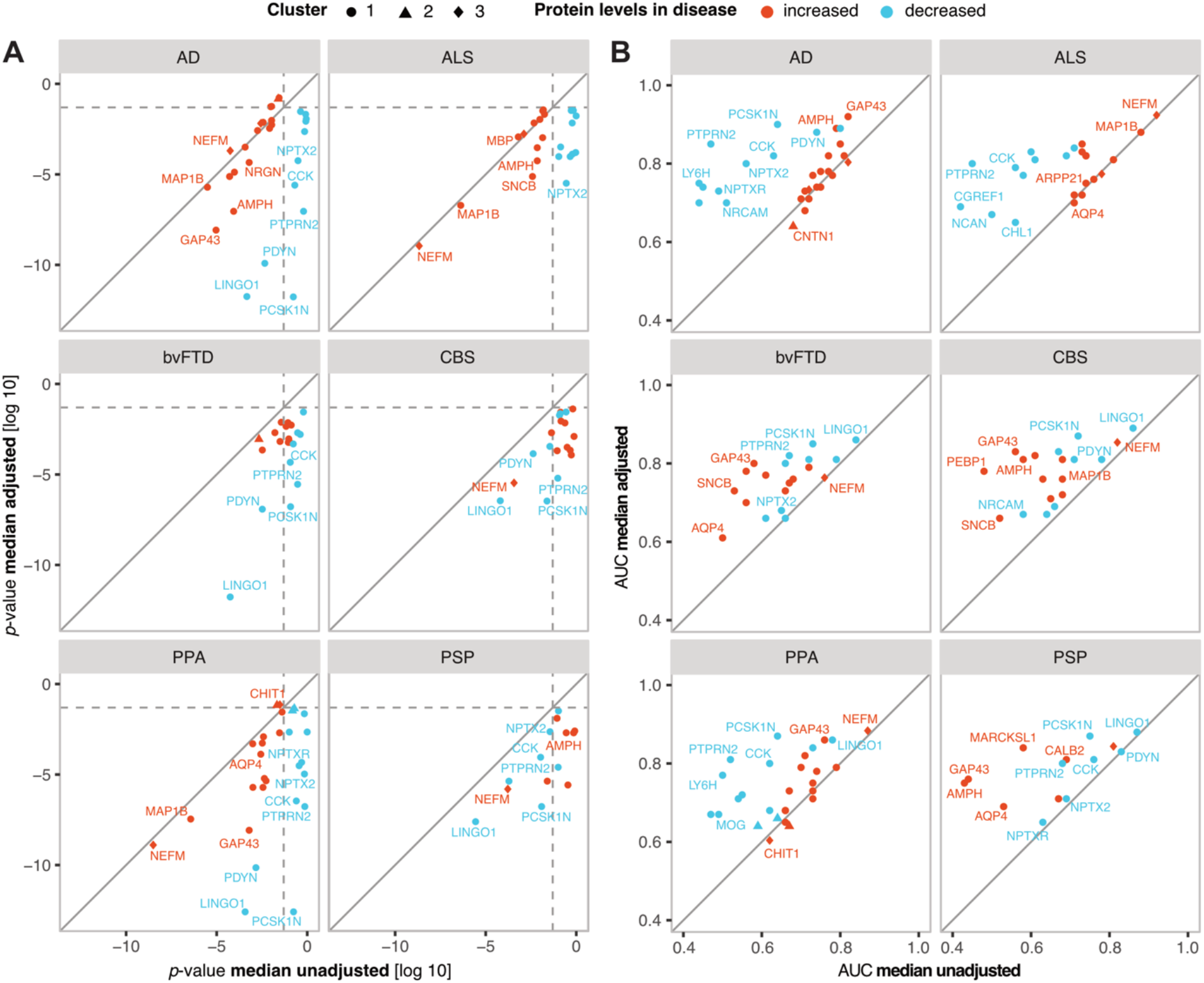
Association with disease and predictive accuracy of individual CSF proteins increases when adjusted for median CSF protein levels. **(A)** A linear model was constructed for each protein and disease, where the protein levels were the outcome variable. In the figure, the p-values for the diagnosis predictor (each disease vs. healthy control) are compared for models adjusted (y-axis) and unadjusted (x-axis) for median CSF protein levels. Only proteins with significant p-value in at least one of the models are included in the figure (**B**) ROC-AUCs resulting from linear models predicting diagnosis (each disease vs. healthy controls) using the adjusted protein levels (y-axis) compared to unadjusted protein levels (x-axis). Only proteins with significant p-value in at least one of the models in (A) are included.

Next, we compared the predictive accuracy for disease vs. healthy controls adjusted and unadjusted individual proteins using logistic regression models, with AUC as the selected metrics. As anticipated, we observed improved performance for the majority of the proteins when adjusted for the median CSF protein levels (**Fig. 2B**).

### The majority of altered proteins are not disease specific

We further investigated the differences in protein levels across the studied diseases. For this, we constructed linear models for comparison between each disease pair, with adjustment for median CSF protein levels, sex and age. As presented in **Fig. 3**, only a handful of proteins showed significant differences in CSF levels between the diseases. Among these proteins, the majority was increased in AD compared to other diseases, with neurogranin (NRGN) showing the highest specificity for AD. Significant differences were also observed for myelin-binding protein (MBP) and neurofilament medium (NEFM), which both showed increased levels in ALS compared to other diseases. However, it is important to note that NEFM also demonstrated higher levels in all diseases compared to healthy controls, indicating a general increase in all the diseases but with larger difference in ALS. Similar results were also obtained from the logistic regression models evaluating the predictability of each measured protein (adjusted for median CSF protein levels) for individual disease against all the other diseases. Based on these outcomes, the majority of the proteins showed low disease specificity (**Supp. Fig. 2**), with only NEFM, NRGN and MBP reaching AUC > 0.7 for one of the diseases versus the others at the same time as for the disease versus healthy controls (**Table 2**).

**Figure 3:**
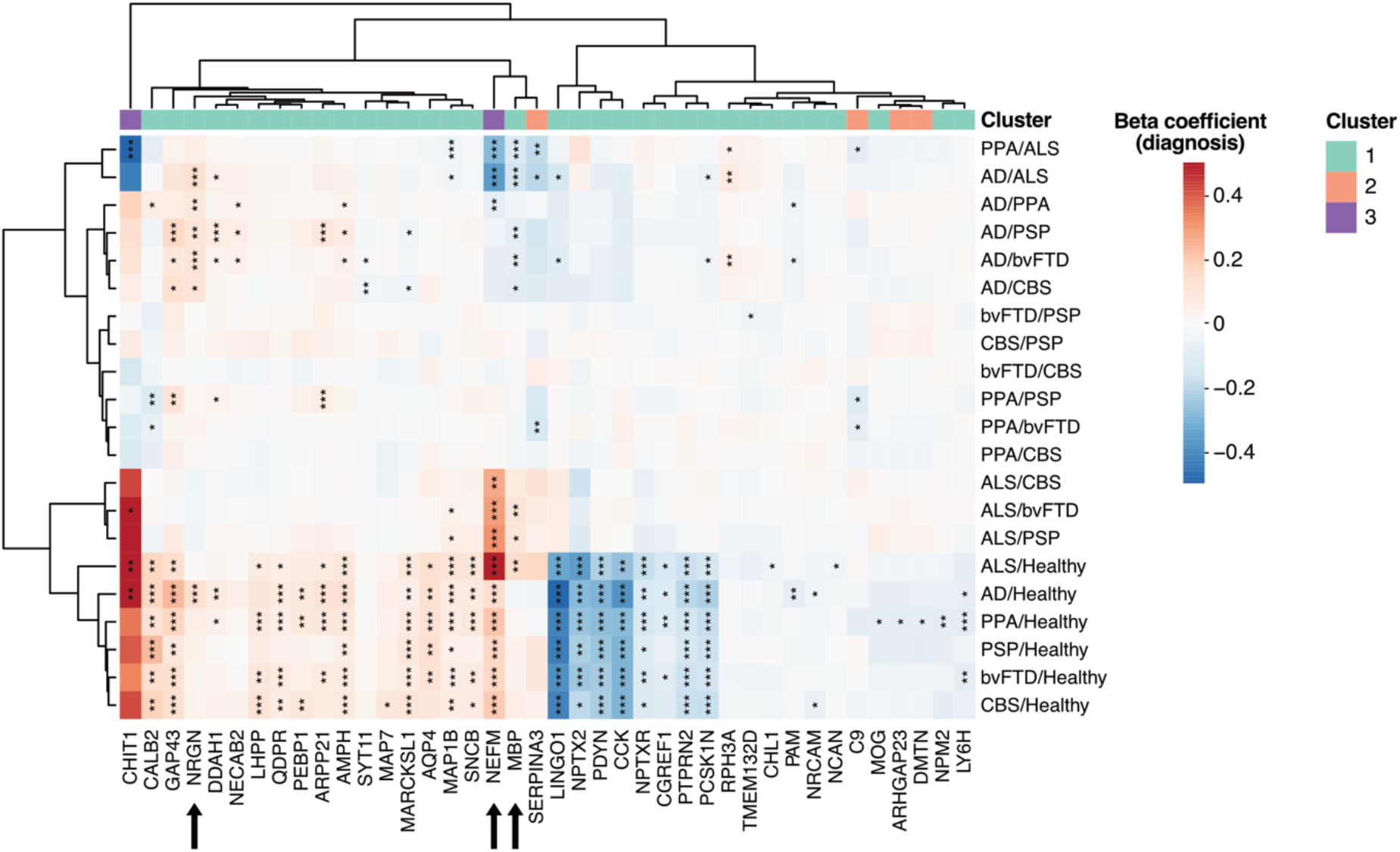
Protein significantly altered in individual diseases compared to healthy controls and/or between the diseases. Heatmap constructed using the beta coefficients for diagnosis from linear models adjusted for median CSF protein levels. Beta coefficients exceeding an absolute value of 0.5 were constrained to 0.5 for visualization purposes. The stars indicate the significance level measured with the p-value: * < 0.05; ** < 0.01, *** < 0.001. Only proteins with significant difference in at least one of the comparisons are included. The proteins are annotated with the clustering results based on protein correlation (Fig. 1A).

### Protein pairs adjust for median CSF protein levels and enhance classification performance

While adjusting for median CSF protein levels enhanced the performance of CSF proteins, it may not be a practical approach. This is particularly true for research studies or in clinical settings where only one or a limited number of proteins is measured, making the estimation of the median CSF protein levels unfeasible. Instead, correlated proteins can be used in pairs to adjust for inter-individual variability. Moreover, proteins in the pair with opposite alteration in disease may further increase the diagnostic performance. To test this, we selected the top five elevated and decreased proteins per disease with the best performance in separating the affected individuals from healthy controls when adjusted for median CSF protein levels (**Supp. Table 3, Supp. Fig. 3**). All protein pair combinations within the 10 selected proteins per disease where then evaluated using logistic models, with the AUC as the evaluation metrics. For all the six investigated diseases, the protein pair combinations with one protein elevated and one decreased in the investigated disease showed the best performance, which was either comparable or higher compared to the performance of the individual proteins adjusted for median CSF protein levels (**Fig. 4, Supp. Fig. 4, Supp. Fig. 5**).

**Figure 4:**
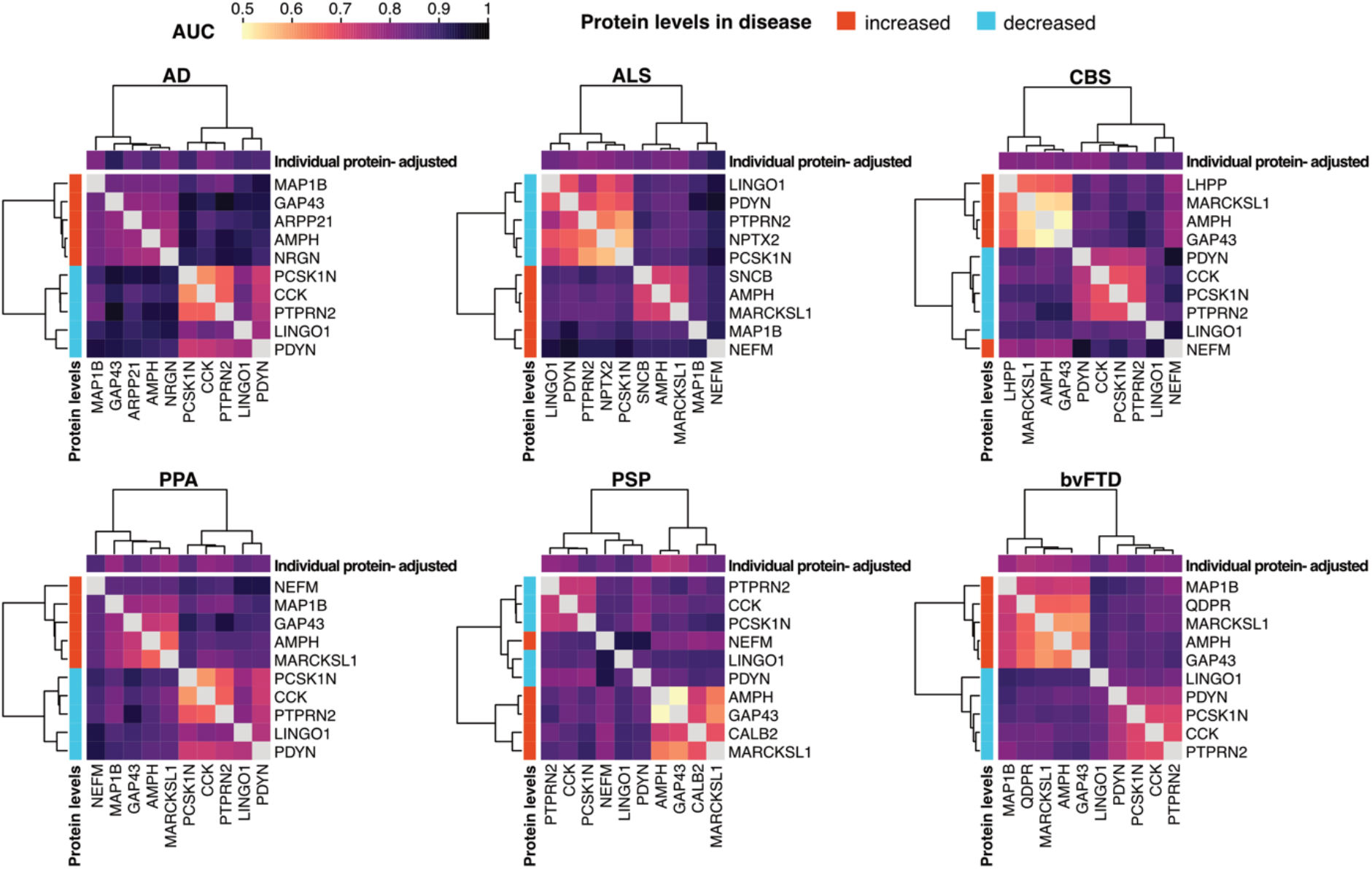
Diagnostic accuracy provided by paired CSF proteins. Heatmaps constructed with AUC values from logistic models predicting diagnosis (each disease vs. healthy control) using protein pairs. For each disease, the selected top five increased and top five decreased proteins (adjusted levels) with the best individual performance in separating the affected individuals from healthy controls were included in the models. Each heatmap is annotated on the left with the direction of the alteration in disease (increased/decreased), and on top with AUC values from logistic models predicting diagnosis (individual disease vs. healthy control) using single proteins adjusted for median CSF levels. AUC values < 0.5 are replaced with the value of 0.5.

**Figure 5:**
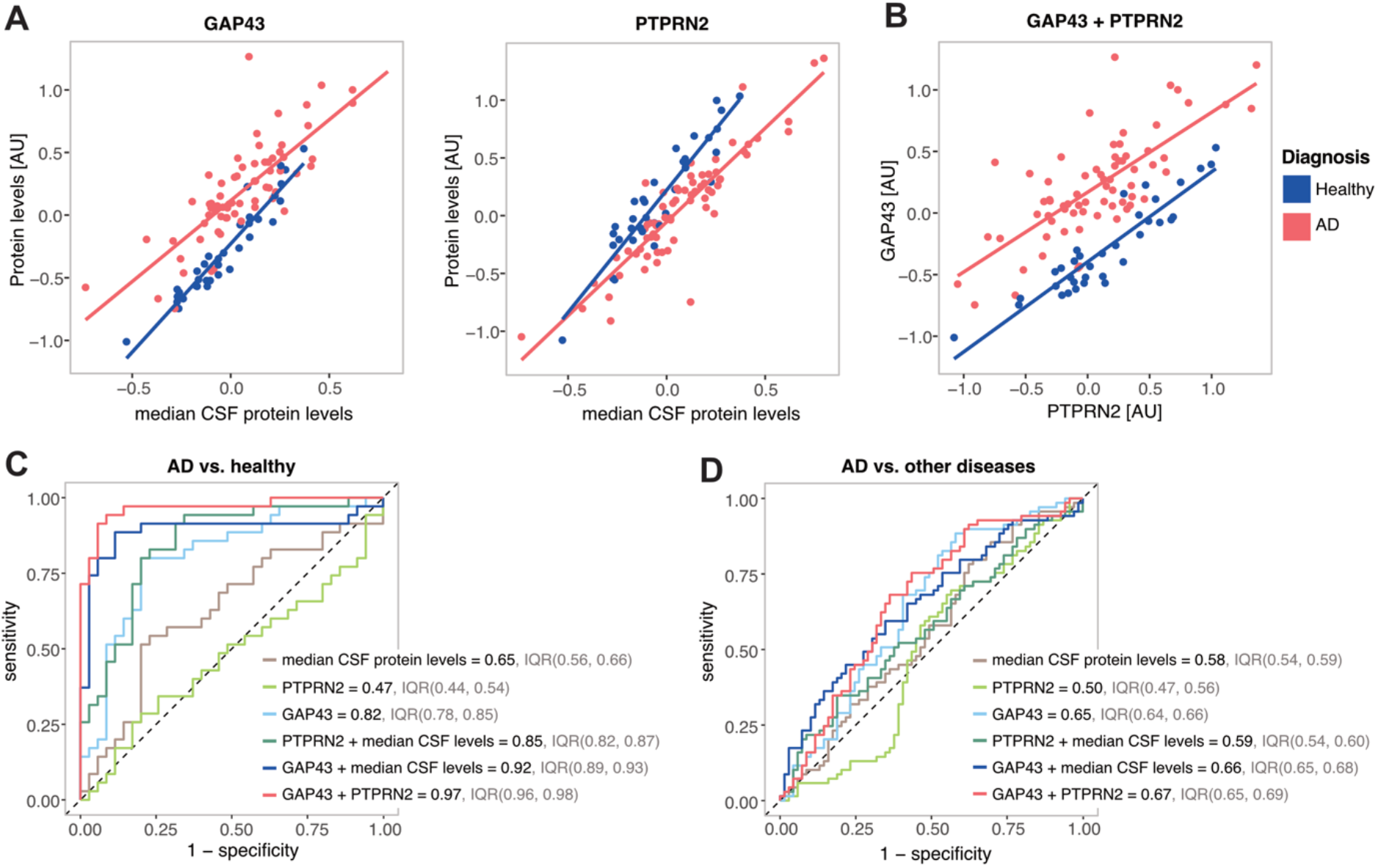
Relationship between GAP43 and PTPRN2 and their diagnostic performance. Examples for AD and healthy controls: (**A**) Scatterplots illustrating the relationship between median CSF protein levels and GAP43 and PTPRN2 levels respectively; (**B**) Scatterplot illustrating the relationship between GAP43 and PTPRN2 levels; (**C, D**) Examples of ROC-curves showing the predictive performance for AD vs. Healthy controls (C), and for AD vs. other diseases (D), using median CSF protein levels, GAP43 and PTPRN2 levels individually, and their combinations.

Furthermore, in ALS, a high performance was observed for all protein combinations including NEFM, suggesting that in this case NEFM is the main contributor to the separation.

To better illustrate the adjustment and the additive effect of the protein pairs, we selected growth associated protein 43 (GAP43) with protein tyrosine phosphatase receptor type N2 (PTPRN2), which provided the best result in separating AD from healthy controls, as an example. **Fig. 5A** shows the separation of AD individuals from healthy controls provided by GAP43 and PTPRN2 individually, when adjusted for median CSF protein levels. **Fig. 5B** further shows the separation provided by GAP43 and PTPRN2 combined. Presented altogether, the performance of the individual and combined components for the AD vs. healthy controls classification using median CSF protein levels, GAP43 and PTPRN2 is compared in **Fig 5C**. The median CSF protein levels, PTPRN2 and GAP43 alone showed the lowest performance (AUC = 0.65, 0.47 and 0.82, respectively). In comparison, PTPRN2 and GAP43 individually but adjusted for median CSF protein levels showed considerably increased performance (AUC = 0.85 and 0.92, respectively). Lastly, GAP3 with PTPRN2 in pair demonstrated the best performance with AUC = 0.97. However, the pair was not able to separate AD from the other diseases (AUC = 0.67), although its performance was better compared to the alternatives (**Fig. 5D**).

### CSF amyloid and tau correlate with median CSF protein levels

Many of the proteins investigated in this study have been previously shown to correlate with the established AD CSF amyloid and tau markers [11, 13-18], suggesting that the established markers may also be affected by the inter-individual variability in general CSF protein levels. All samples in this cohort, except healthy controls, were analysed for Aβ40, Aβ42, p-tau and t-tau (**Table 1**). All four markers showed correlation to median CSF protein levels in this cohort (**Fig. 6A)**. Notably, the strongest correlation was observed between the median CSF protein levels and Aβ40 (rho = 0.74 for all samples). Contrary to Aβ42, CSF p-tau and t-tau are not commonly adjusted for general CSF protein levels. Therefore, individuals with not only high CSF tau levels but also high general CSF protein levels could be falsely denotated as CSF-tau positive based on the set clinical cut-offs. To explore whether this could be the case, we compared the distribution of the median CSF protein levels in sample group categorised based on the CSF Aβ42/40 and p-tau status (**Supp. Table 4**). As anticipated, the amyloid-negative but tau-positive individuals (A-T+) showed generally higher CSF protein levels compared to all the other groups (**Fig. 6B**), indicating a possible false p-tau-positivity derived from overall high CSF protein levels in this group.

**Figure 6:**
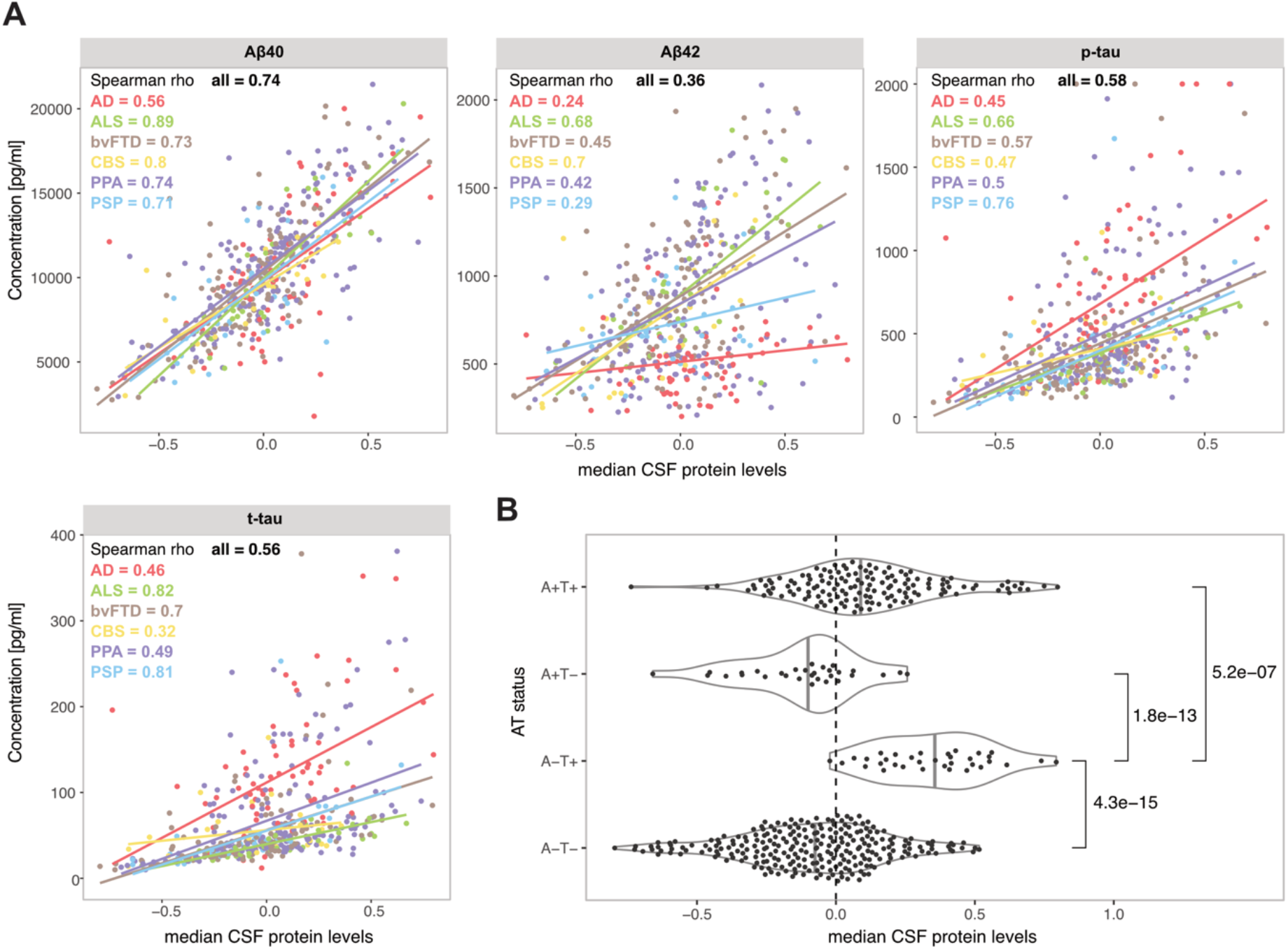
Relationship between median CSF protein levels and CSF amyloid and tau. **(A**) Scatterplots illustrating the relationship between Aβ40, Aβ42, p-tau and t-tau and median CSF protein levels for all individuals as well as for individual diseases. (**B**) Distribution of median CSF protein levels in groups of participants divided by amyloid and tau (AT) status determined with CSF Aβ42/Aβ40 and p-tau levels.

## Discussion

The effectiveness of a biomarker often relies on an alteration in its levels from a healthy state. However, this change might be relatively small compared to the naturally occurring variability among individuals, which makes the detection of pathology-related protein alterations challenging. In this study, we show that CSF levels of brain-derived proteins co-vary with their median level (here referred to as median CSF protein levels), which represent the main source of variability in the brain derived proteins between individuals. We further demonstrate that adjusting for the median levels increases the association of the individual proteins with disease as well as improve their performance in distinguishing individuals with disease from healthy controls. In addition, we show that this adjustment can be simplified by combining proteins in pairs, where one of the proteins increases and one decreases in the studied disease. Using this approach, we achieved adjustment for individual-dependent overall protein levels and further increased the classification performance through the additive effect of the two proteins with opposite alteration.

Based on the data in this cohort of 499 individuals, the median CSF levels of brain derived proteins did not significantly differ between males and females. However, weak but significant positive correlation was detected between the CSF median levels and age in this cohort. This is in concordance with previous studies indicating higher levels of many CSF proteins in older individuals, possibly due to decreased CSF turnover in older age [33-35]. We also observed significant differences in median CSF protein levels between the diagnostic groups, with slightly higher levels in AD, ALS and PPA. This subtle difference may stem from the limited protein selection, wherein many proteins exhibit more pronounced increases in these diseases, consequently slightly elevating the overall group median. Alternatively, variations in CSF physiology across the groups could contribute [36]. However, these observed group differences were modest, with considerable overlap in group variance, leading us to suggest that inter-individual variability in median CSF protein levels is not mainly influenced by disease pathology.

Consistent with our hypothesis that inter-individual variability in general levels of brain-derived proteins partially masks the variability driven by pathological changes, we found that when adjusted for median CSF protein levels, majority of the measured protein strengthened their association with disease. This pattern was evident for proteins both increased and decreased in disease across all the studied groups. Several proteins were consistently altered across all the diseases. Leucine rich repeat and Ig domain containing 1 protein (LINGO1), neuronal pentraxin 2 (NPTX2), prodynorphin (PDYN), cholecystokinin (CCK), neuronal pentraxin receptor (NPTXR), PTPRN2 and proprotein convertase subtilisin/kexin type 1 inhibitor (PCSK1N) were found significantly decreased in all six diseases compared to healthy controls. Previous studies have also reported reductions in these proteins in CSF across various neurodegenerative disorders [28, 37-45]. Importantly, in our study cohort, most of these alterations reached statistical significance only after adjustment for median CSF protein levels. Conversely, NEFM, GAP43, amphiphysin (AMPH), myristoylated alanine-rich C-kinase substrate like 1 (MARCKSL1) and calbindin 2 (CALB2) showed consistent increase in all six diseases. Among these, NEFM, GAP43 and AMPH have previously been identified as elevated in CSF in association with neurodegeneration [13, 17, 25, 28, 46, 47]. Like GAP43, MARCKSL1 is a substrate of protein kinase C and is suggested to play a role in the regulation of pre- and post-synapse morphologies [48, 49]. The increase in CSF levels of MARCKSL1 and GAP43 may thus be related. CALB2, a calcium-binding protein highly expressed in neurons, has previously been found elevated in CSF from individuals with Niemann-Pick disease, along with NEFL and tau [50-52], suggesting a potential association to neuronal damage. The considerable overlap in altered proteins across the studied diseases indicates that changes in the levels of many CSF proteins may stem from common mechanisms to the diseases, potentially reflecting general neurodegenerative processes and/or neuronal loss. While such biomarkers may offer limited utility for differential diagnostics, they hold promise as valuable tools for monitoring disease progression and assessing treatment efficacy.

When comparing CSF protein patterns across the six diseases, only three proteins exhibited some degree of specificity toward a particular disease when adjusted for median CSF protein levels. NRGN was found to be elevated in AD compared to all other diseases and healthy controls. This observation aligns with previous cross-disease investigations conducted by Wellington *et al*. [53] and Portelius *et al*. [54], where NRGN levels were elevated in AD but not in FTD, Lewy body dementia, Parkinson’s disease, multiple system atrophy, PSP, PPA, ALS, or CBS. Further, we observed MBP to be significantly increased in ALS compared to the other diseases, except CBS (potentially due to low number of individuals in this group), as well as compared to healthy controls. MBP, an abundant component of the myelin membrane, is considered to be a nonspecific marker of CNS inflammation [55], and has previously been studied as a potential biomarker for multiple sclerosis, showing elevated CSF levels in the disease [56, 57]. Recently, Yue Li *et al*. demonstrated an increase in CSF MBP in 16% of patients with ALS, with a higher increase in those with comorbid bvFTD [58]. In our study, we did not observe any significance difference between the included ALS subgroups (ALS, ALS + bvFTD, ALS+PPA, data not shown). However, it is important to note that both studies had limited participant numbers, thus further research is needed to determine the significance of MBP as biomarker in the context of ALS. Lastly, NEFM showed significant elevation in ALS compared to the other diseases. However, NEFM was also significantly elevated in all the diseases compared to other controls, as discussed earlier, indicating a non-specific increase in neurodegenerative conditions, with the most pronounced increase in ALS. This pattern aligns with the observations for NEFL [59], suggesting that both reflect similar processes, although NEFM is not as extensively studied.

We further suggest an alternative method for addressing inter-individual variance in general CSF levels of brain-derived proteins by utilizing proteins in pairs. This approach is particularly valuable for implementing biomarkers in clinical practice, where precise and reliable procedures are essential, and the number of measured proteins is limited. Furthermore, it allows for further increase in predictive accuracy through additive effect when correlated proteins with opposite alterations in CSF are combined. Building upon our previous work focused on AD [19], we demonstrate here the applicability of this adjustment method in the broader context of neurodegenerative diseases. Additionally, given our findings suggesting that overall CSF levels of brain-derived proteins are not primarily driven by pathology, we propose that this adjustment could be beneficial also for CSF protein studies in other CNS-associated research fields, such as in biomarker studies within neuro-inflammatory and psychiatric disorders. The use of biomarker ratios is not a novel concept. For instance, commonly employed protein ratio in both research and in clinical practice is that of CSF Aβ42 to Aβ40, a method used to determine the presence of amyloid pathology. In this context, Aβ40 is considered an adjustment factor correcting for the individual-specific variability in amyloid production levels [10]. Here we show that Aβ40 strongly correlates with the median CSF levels of brain derived proteins, indicating that Aβ40 may work not only as an adjustment factor for amyloid production but also for the overall CSF protein levels derived from the individual-specific CSF dynamics. Notably, we also observed a moderately strong correlation between median CSF protein levels and p-tau and t-tau, suggesting that these markers are likely also affected by the interindividual variability in the overall CSF protein levels and could thus benefit from adjustment. This hypothesis is supported by previous work reported by Guo *et al*. who demonstrated that CSF adjusting p-tau181 with Aβ40 enhances predictive accuracy for the presence of tauopathy compared to p-tau181 alone [60]. Overall high CSF concentration of brain-derived proteins could also explain the existence of a clinically controversial group of individuals with amyloid-negative but tau-positive status as determined based on CSF Aβ42/Aβ40 and p-tau. According to our data, individuals in this group have considerably higher median CSF protein levels compared to the other diagnostic groups, suggesting that elevated tau levels in this group may not necessarily denote tau pathology but rather be a result of impaired CSF dynamics causing overall elevation in CSF protein concentrations, as also suggested by Eriksson *et al*. [61] and Karlsson *et al*. [62].

This study has several limitations that require consideration. One constraint is the relatively small number of measured proteins, which are pre-selected and may not fully represent the CSF protein repertoire. This partial representation could potentially impact the accuracy of the determined median CSF levels of brain-derived proteins. Due to the limited number of healthy controls, the protein clustering is performed based on correlations between the proteins constructed from the full sample cohort. This might lead to weaker correlations for proteins that alter with pathology. Further limiting is the lack of direct validation in an independent cohort, which was not available for this study. Although our findings are well supported by previous research, further validation would strengthen our conclusions. Additionally, while the study cohort is relatively large, the number of participants in some of the diagnostic groups was small, limiting the statistical power of the performed comparisons. Further, given the cross-sectional nature of this study, the identified proteins and protein pairs cannot be evaluated as prospective biomarkers. To assess their potential for monitoring disease progression, further investigation using longitudinal samples and clinical follow-up is necessary. Finally, it would be valuable to compare the adjusted protein data with other types of biomarkers such as MRI and/or PET images to determine their concordance.

## Conclusion

In this study we show that inter-individual variability in the overall CSF levels of brain-derived proteins partially masks the association of single proteins with disease pathology, and that adjusting for this variability enhances their predictive accuracy. Furthermore, we simplify this adjustment by using correlated proteins in pairs where they adjust for each other and further enhance the performance when having opposite alteration in disease. Importantly, as p-tau and t-tau also demonstrate correlation to the median CSF levels of brain-derived proteins, we suggest that these established biomarkers could also benefit from such adjustment. Upon closer investigation of the adjusted proteins measured in this study across several neurodegenerative diseases, we find that many CSF proteins are similarly altered in all or majority of the studied diseases, indicating that these proteins likely reflect common neuro-degenerative processes. This highlights the importance of cross-disease study design also for future studies to accurately determine the specificity and clinical applicability of individual biomarkers.

## Data Availability

All data produced in the present study are available upon reasonable request to the authors

## List of abbreviations

AD: Alzheimer’s disease
ALS: amyotrophic lateral sclerosis
AMPH: amphiphysin
AUC: area under the curve
Aβ40: amyloid beta 40 residues long
Aβ42: amyloid beta 42 residues long
bvFTD: behavioural variant frontotemporal dementia
CALB2: calbindin 2
CBS: corticobasal syndrome
CCK: cholecystokinin
CSF: cerebrospinal fluid
GAP43: growth associated protein 43
LINGO1: leucine rich repeat and Ig domain containing 1
MARCKSL1: myristoylated alanine-rich C-kinase substrate like 1
MBP: myelin-binding protein
MND: motor neurone disease
NEFL: neurofilament light chain
NEFM: neurofilament medium chain
NPTX2: neuronal pentraxin 2
NPTXR: neuronal pentraxin receptor
NRGN: neurogranin
p-tau: phosphorylated tau
PCA: principal component analysis
PCSK1N: proprotein convertase subtilisin/kexin type 1 inhibitor
PDYN: prodynorphin
PPA: primary progressive aphasia
PSP: progressive supranuclear palsy
PTPRN2: protein tyrosine phosphatase receptor type N2
Q-Alb: albumin CSF/serum quotient
ROC: receiver operating characteristic
t-tau: total tau

## Declarations

### Ethical approval and consent to participate

The study was conducted following the Declaration of Helsinki and approved by the local ethics committees of all centres involved (University of Ulm #39/11). An additional ethical approval for the presented study was received from the Swedish Ethical Review Authority (2022-02653-01). All participants in both cohorts gave written informed consent for sample and data collection.

### Availability of data and materials

The datasets used and/or analysed during the current study are available from the corresponding author on reasonable request.

## Acknowledgement

We express our gratitude to all participants and their families for contributing to the study. We also thank to all members of the German FTLD consortium and to the staff involved in the Human Protein Atlas for their efforts.

## Author’s contributions

SM, PN and AM designed the study and SM performed the experimental work. SM performed the data analysis, visualisations and data interpretations with support from SB, AM, PN, JO, NG, and MO. SAS, JL, JDS, KFa, KFl, HJ, JK, BL, ML, ACL, JP, AS, MLS, JW, PS and MO organised the collection of samples and clinical data from the recruited participants. PN and AM supervised the project. SM wrote the manuscript with input from SB, PN, AM, JO and MO. All authors approved the manuscript.

