## Supplementary material for "Addressing inter-individual variability in CSF levels of brain-derived proteins across neurodegenerative diseases"

### Supplementary figures and tables

Supplementary figures are listed first, followed by supplementary tables.

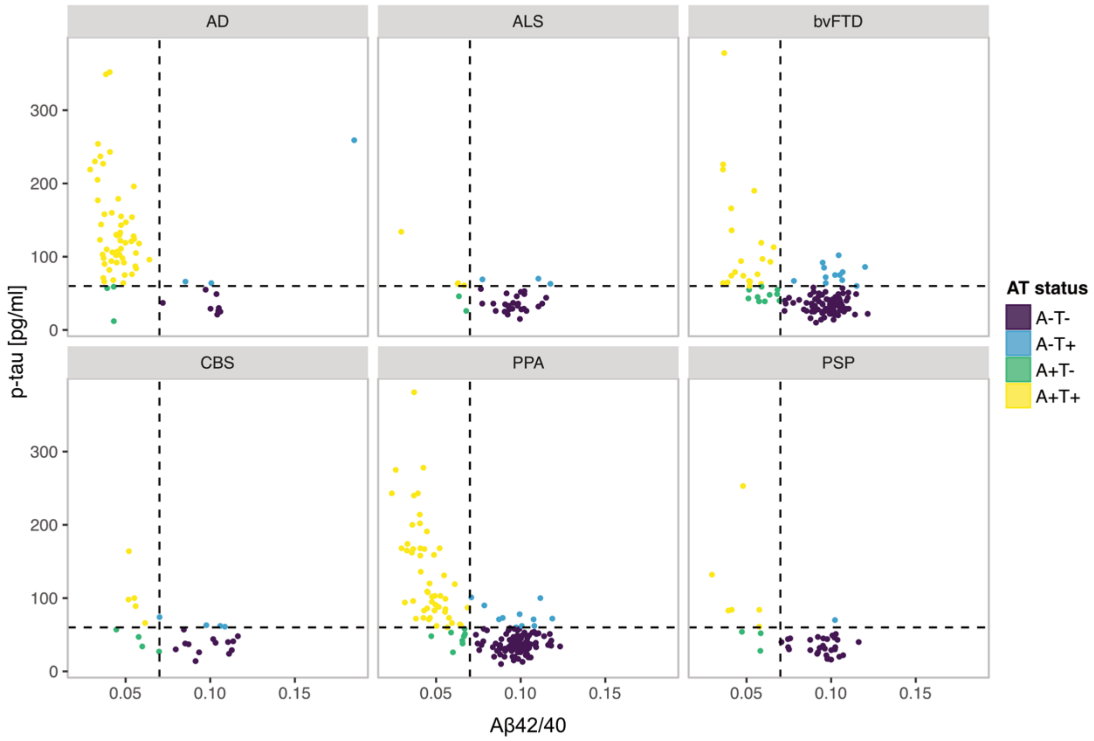


**Supplementary figure 1:** CSF Aβ42/40 and p-tau levels per diagnosis. The dashed lines label the classification cut-offs: Aβ42/40 = 0.07; p-tau = 60 pg/ml.

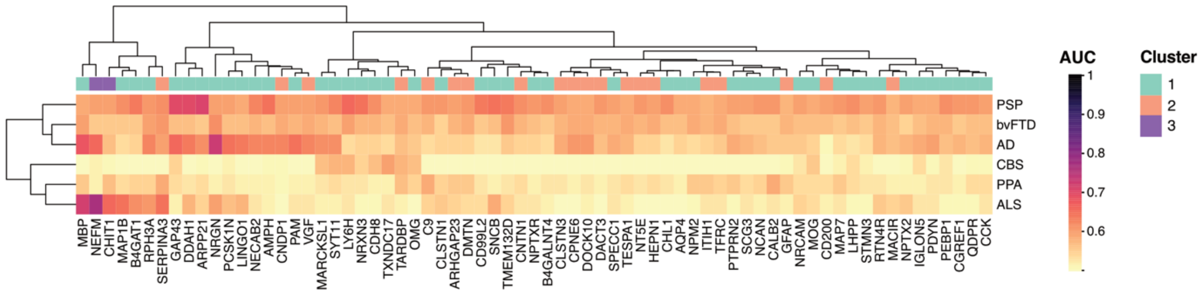


**Supplementary figure 2:** Disease specificity of CSF proteins. A heatmap showing predictive accuracy (evaluated with AUC) of individual proteins for each disease vs. all diseases, with adjustment for median CSF protein levels. The predicted disease is indicated on the right. The heatmap is annotated on top with the clustering results based on protein correlation (Fig. 1A).

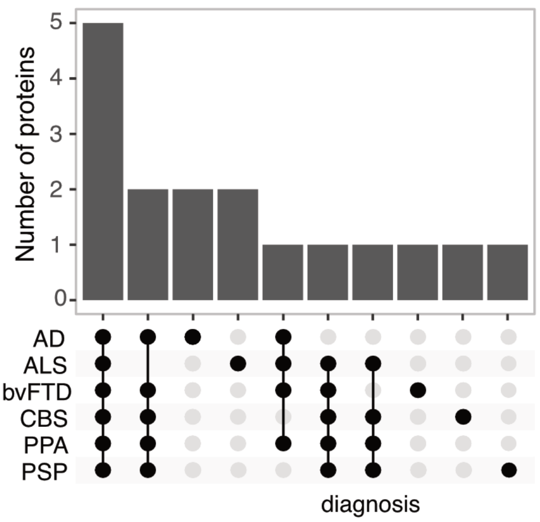


**Supplementary figure 3:** Top 10 proteins with best performance in classifying disease vs. healthy controls – overlap between diseases

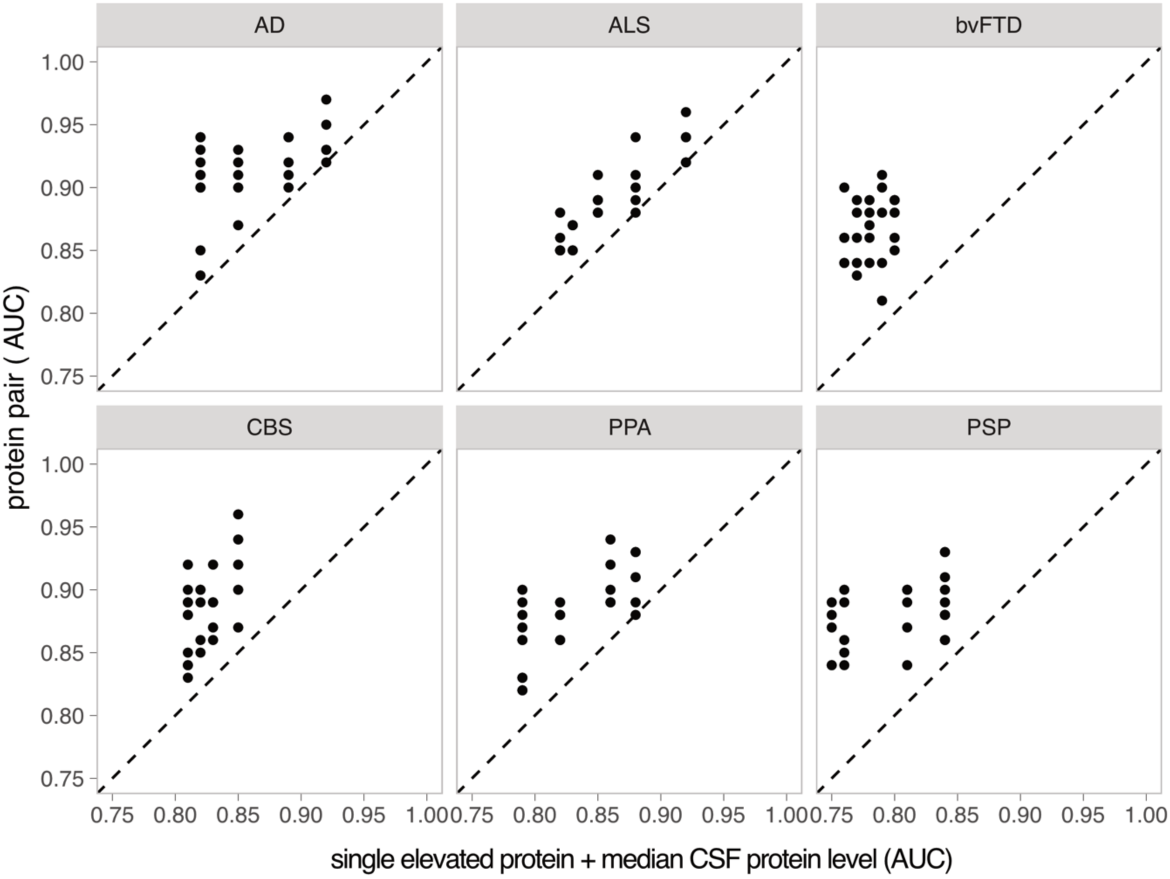


**Supplementary figure 4:** AUC obtained from logistic regression models for disease versus healthy controls classification. The x-axis represents the AUC values obtained from models with single proteins *elevated* in disease and CSF median protein levels as predictors. On the y-axis, AUC values are depicted for models with protein pairs, where one protein is elevated and one is decreased in disease, as predictors.

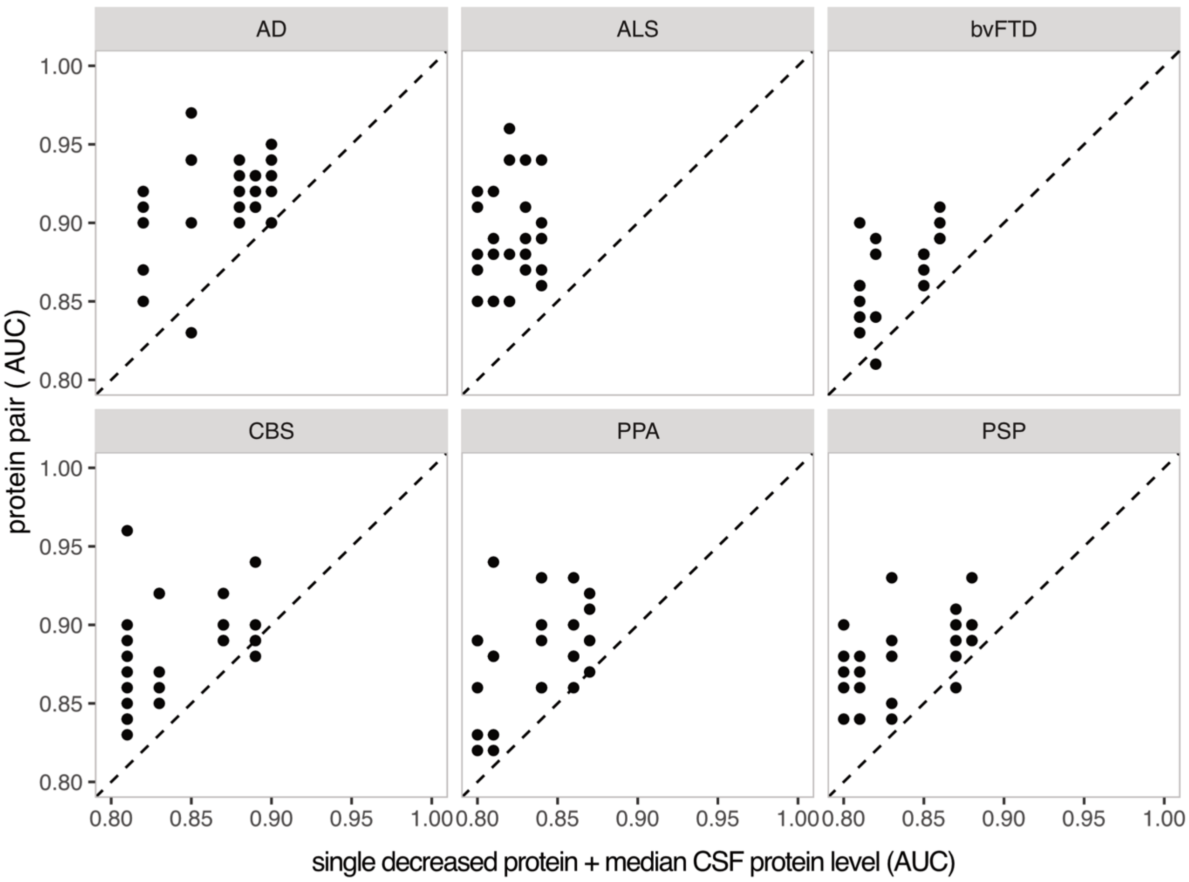


**Supplementary figure 5:** AUC obtained from logistic regression models for disease versus healthy controls classification. The x-axis represents the AUC values obtained from models with single proteins *decreased* in disease and CSF median protein levels as predictors. On the y-axis, AUC values are depicted for models with protein pairs, where one protein is elevated and one is decreased in disease, as predictors.

**Supplementary table 1:** List of analysed proteins and used antibodies.

| HGNC ID | Protein name | Uniprot ID | Antibody | Sample dilution |
|---|---|---|---|---|
| **AMPH** | Amphiphysin | P49418 | HPA019829 | 1:25 |
| **AQP4** | Aquaporin 4 | P55087 | HPA014784 | 1:200 |
| **ARHGAP23** | Rho GTPase activating protein 23 | Q9P227 | HPA019818 | 1:25 |
| **ARPP21** | CAMP regulated phosphoprotein 21 | Q9UBL0 | HPA017303 | 1:25 |
| **B4GALNT4** | Beta-1,4-N-acetyl-galactosaminyltransferase 4 | Q76KP1 | HPA053126 | 1:25 |
| **B4GAT1** | Beta-1,4-glucuronyltransferase 1 | O43505 | HPA015484 | 1:200 |
| **C9** | Complement C9 | P02748 | HPA070709 | 1:25 |
| **CALB2** | Calbindin 2 | P22676 | HPA007305 | 1:25 |
| **CCK** | Cholecystokinin | P06307 | HPA069515 | 1:200 |
| **CD200** | CD200 molecule | P41217 | HPA031149 | 1:25 |
| **CD99L2** | CD99 molecule like 2 | Q8TCZ2 | HPA061400 | 1:200 |
| **CDH8** | Cadherin 8 | P55286 | HPA014908 | 1:200 |
| **CGREF1** | Cell growth regulator with EF-hand domain 1 | Q99674 | HPA008241 | 1:200 |
| **CHIT1** | Chitinase 1 | Q13231 | HPA010575 | 1:25 |
| **CHL1** | Cell adhesion molecule L1 like | O00533 | HPA003345 | 1:200 |
| **CLSTN1** | Calsyntenin 1 | O94985 | HPA012412 | 1:200 |
| **CLSTN3** | Calsyntenin 3 | Q9BQT9 | HPA070830 | 1:200 |
| **CNDP1** | Carnosine dipeptidase 1 | Q96KN2 | HPA016933 | 1:200 |
| **CNTN1** | Contactin 1 | Q12860 | HPA070467 | 1:25 |
| **CPNE6** | Copine 6 | O95741 | HPA031636 | 1:200 |
| **DACT3** | Dishevelled binding antagonist of beta catenin 3 | Q96B18 | HPA043053 | 1:200 |
| **DDAH1** | Dimethylarginine dimethylaminohydrolase 1 | O94760 | HPA006308 | 1:25 |
| **DMTN** | Dematin actin binding protein | Q08495 | HPA024290 | 1:200 |
| **DOCK10** | Dedicator of cytokinesis 10 | Q96BY6 | HPA058106 | 1:200 |
| **GAP43** | Growth associated protein 43 | P17677 | HPA013603 | 1:200 |
| **GFAP** | Glial fibrillary acidic protein | P14136 | 16825-1-AP | 1:25 |
| **HEPN1** | Hepatocellular carcinoma, down-regulated 1 | Q6WQI6 | HPA063054 | 1:25 |
| **IGLON5** | IgLON family member 5 | A6NGN9 | HPA041994 | 1:25 |
| **ITIH1** | Inter-alpha-trypsin inhibitor heavy chain 1 | P19827 | HPA042049 | 1:200 |
| **LHPP** | Phospholysine phosphohistidine inorganic pyrophosphate phosphatase | Q9H008 | HPA009269 | 1:25 |
| **LINGO1** | Leucine rich repeat and Ig domain containing 1 | Q96FE5 | HPA074653 | 1:200 |
| **LY6H** | Lymphocyte antigen 6 family member H | O94772 | HPA077218 | 1:25 |
| **MACIR** | Macrophage immunometabolism regulator | Q96GV9 | HPA043434 | 1:25 |
| **MAP1B** | Microtubule associated protein 1B | P46821 | HPA022275 | 1:25 |
| **MAP7** | Microtubule associated protein 7 | Q14244 | HPA029712 | 1:25 |
| **MARCKSL1** | MARCKS like 1 | P49006 | HPA030528 | 1:25 |
| **MBP** | Myelin basic protein | P02686 | HPA049222 | 1:25 |
| **MOG** | Myelin oligodendrocyte glycoprotein | Q16653 | HPA021873 | 1:200 |
| **NCAN** | Neurocan | O14594 | HPA058000 | 1:200 |
| **NECAB2** | N-terminal EF-hand calcium binding protein 2 | Q7Z6G3 | HPA013998 | 1:25 |
| **NEFM** | Neurofilament medium | P07197 | HPA022845 | 1:25 |
| **NPM2** | Nucleophosmin/nucleoplasmin 2 | Q86SE8 | HPA041070 | 1:25 |
| **NPTX2** | Neuronal pentraxin 2 | P47972 | HPA058320 | 1:200 |
| **NPTXR** | Neuronal pentraxin receptor | O95502 | HPA001079 | 1:25 |
| **NRCAM** | Neuronal cell adhesion molecule | Q92823 | HPA061433 | 1:25 |

| HGNC ID | Protein name | Uniprot ID | Antibody | Sample dilution |
|---|---|---|---|---|
| **NRGN** | Neurogranin | Q92686 | HPA038171 | 1:25 |
| **NRXN3** | Neurexin 3 | Q9HDB5, Q9Y4C0 | HPA002727 | 1:200 |
| **NT5E** | 5'-nucleotidase ecto | P21589 | HPA048043 | 1:25 |
| **OMG** | Oligodendrocyte myelin glycoprotein | P23515 | HPA008206 | 1:200 |
| **PAM** | Peptidylglycine alpha-amidating monooxygenase | P19021 | HPA042260 | 1:200 |
| **PCSK1N** | Proprotein convertase subtilisin/kexin type 1 inhibitor | Q9UHG2 | HPA064734 | 1:200 |
| **PDYN** | Prodynorphin | P01213 | HPA053342 | 1:200 |
| **PEBP1** | Phosphatidylethanolamine binding protein 1 | P30086 | HPA063904 | 1:200 |
| **PTPRN2** | Protein tyrosine phosphatase receptor type N2 | Q92932 | HPA007255 | 1:200 |
| **QDPR** | Quinoid dihydropteridine reductase | P09417 | HPA058951 | 1:25 |
| **RPH3A** | Rabphilin 3A | Q9Y2J0 | HPA002475 | 1:25 |
| **RTN4R** | Reticulon 4 receptor | Q9BZR6 | HPA063584 | 1:200 |
| **SCG3** | Secretogranin III | Q8WXD2 | HPA053715 | 1:200 |
| **SERPINA3** | Serpin family A member 3 | P01011 | HPA000893 | 1:200 |
| **SNCB** | Synuclein beta | Q16143 | HPA035876 | 1:200 |
| **SPECC1** | Sperm antigen with calponin homology and coiled-coil domains 1 | Q5M775 | HPA021421 | 1:25 |
| **STMN3** | Stathmin 3 | Q9NZ72 | HPA012947 | 1:25 |
| **SYT11** | Synaptotagmin 11 | Q9BT88 | HPA064091 | 1:25 |
| **TARDBP** | TAR DNA binding protein | Q13148 | 10782-2-AP | 1:25 |
| **TESPA1** | Thymocyte expressed, positive selection associated 1 | A2RU30 | HPA058823 | 1:200 |
| **TFRC** | Transferrin receptor | P02786 | HPA028598 | 1:200 |
| **TMEM132D** | Transmembrane protein 132D | Q14C87 | HPA010739 | 1:25 |
| **TXNDC17** | Thioredoxin domain containing 17 | Q9BRA2 | HPA022931 | 1:25 |
| **VGF** | VGF nerve growth factor inducible | O15240 | HPA058371 | 1:200 |

**Supplementary table 2:** *p*-values from Dunn's test comparing median CSF levels across diagnostic groups

| comparison | p-value | adjusted p-value | significant |
|---|---|---|---|
| **AD - ALS** | 0.734 | 0.856 | no |
| **AD - CBS** | 0.051 | 0.133 | no |
| **AD - Healthy control** | 0.046 | 0.160 | no |
| **AD - PPA** | 0.286 | 0.462 | no |
| **AD - PSP** | 0.003 | 0.059 | no |
| **AD - bvFTD** | 0.003 | 0.030 | yes |
| **ALS - CBS** | 0.143 | 0.334 | no |
| **ALS - Healthy control** | 0.150 | 0.316 | no |
| **ALS - PPA** | 0.658 | 0.813 | no |
| **ALS - PSP** | 0.023 | 0.098 | no |
| **ALS - bvFTD** | 0.049 | 0.148 | no |
| **CBS - Healthy control** | 0.892 | 0.937 | no |
| **CBS - PPA** | 0.159 | 0.304 | no |
| **CBS - PSP** | 0.557 | 0.731 | no |
| **Healthy control - PPA** | 0.160 | 0.279 | no |
| **Healthy control - PSP** | 0.429 | 0.601 | no |
| **PPA - PSP** | 0.012 | 0.086 | no |
| **bvFTD - CBS** | 0.983 | 0.983 | no |
| **bvFTD - Healthy control** | 0.872 | 0.964 | no |
| **bvFTD - PPA** | 0.013 | 0.067 | no |
| **bvFTD - PSP** | 0.401 | 0.602 | no |

**Supplementary table 3:** Top five elevated and decreased proteins per disease with the best performance in separating the affected individuals from healthy controls when adjusted for median CSF protein levels.

| | **AD** | | **ALS** | | **CBS** | | **PPA** | | **PSP** | | **bvFTD** | |
|---|---|---|---|---|---|---|---|---|---|---|---|---|
| | **protein** | **AUC** | **protein** | **AUC** | **protein** | **AUC** | **protein** | **AUC** | **protein** | **AUC** | **protein** | **AUC** |
| **elevated in disease** | GAP43 | 0.92 | NEFM | 0.92 | NEFM | 0.85 | NEFM | 0.88 | MARCKSL1 | 0.84 | GAP43 | 0.8 |
| | AMPH | 0.89 | MAP1B | 0.88 | GAP43 | 0.83 | GAP43 | 0.86 | NEFM | 0.84 | MAP1B | 0.79 |
| | ARPP21 | 0.85 | SNCB | 0.85 | MARCKSL1 | 0.82 | AMPH | 0.82 | CALB2 | 0.81 | AMPH | 0.78 |
| | MAP1B | 0.82 | AMPH | 0.83 | LHPP | 0.81 | MAP1B | 0.79 | GAP43 | 0.76 | MARCKSL1 | 0.77 |
| | NRGN | 0.82 | MARCKSL1 | 0.82 | AMPH | 0.81 | MARCKSL1 | 0.79 | AMPH | 0.75 | QDPR | 0.76 |
| **decreased in disease** | PCSK1N | 0.9 | LINGO1 | 0.84 | LINGO1 | 0.89 | PCSK1N | 0.87 | LINGO1 | 0.88 | LINGO1 | 0.86 |
| | LINGO1 | 0.89 | PCSK1N | 0.83 | PCSK1N | 0.87 | LINGO1 | 0.86 | PCSK1N | 0.87 | PCSK1N | 0.85 |
| | PDYN | 0.88 | PDYN | 0.82 | PTPRN2 | 0.83 | PDYN | 0.84 | PDYN | 0.83 | PTPRN2 | 0.82 |
| | PTPRN2 | 0.85 | NPTX2 | 0.81 | PDYN | 0.81 | PTPRN2 | 0.81 | CCK | 0.81 | PDYN | 0.81 |
| | CCK | 0.82 | PTPRN2 | 0.8 | CCK | 0.81 | CCK | 0.8 | PTPRN2 | 0.8 | CCK | 0.81 |

**Supplementary table 4:** Number of individuals per AT group and diagnosis.

| | **AT status** | | | |
|---|---|---|---|---|
| **Diagnosis** | A+T+ | A+T- | A-T+ | A-T- |
| AD | 55 | 3 | 3 | 8 |
| ALS | 4 | 2 | 3 | 26 |
| CBS | 5 | 4 | 4 | 13 |
| PPA | 51 | 8 | 11 | 96 |
| PSP | 6 | 3 | 1 | 29 |
| bvFTD | 21 | 10 | 12 | 86 |
| **total** | 142 | 30 | 34 | 258 |
